# Investigating Electrocardiographic Abnormalities in Patients with Coronary Microvascular Dysfunction

**DOI:** 10.1101/2024.04.21.24306154

**Authors:** Ilan Merdler, Andrew P. Hill, Sevket Tolga Ozturk, Matteo Cellamare, Cheng Zhang, Avantika Banerjee, Kalyan R. Chitturi, Lior Lupu, Vaishnavi Sawant, Itsik Ben-Dor, Ron Waksman, Hayder D. Hashim, Brian C. Case

## Abstract

**BACKGROUND:** Coronary microvascular dysfunction (CMD) is a frequently overlooked and underdiagnosed mechanism of cardiac ischemia despite its heightened risk of major adverse cardiovascular events. The potential impact of CMD on the conduction system and occurrence of arrhythmias has received limited attention. This study aimed to examine the link between CMD and surface electrocardiography (ECG).

**METHODS:** We harnessed the Coronary Microvascular Disease registry (CMDR), utilizing the CoroVentis CoroFlow System (Abbott) to invasively assess patients presenting with symptomatic chest pain. All patients underwent microvascular assessment of the left anterior descending artery territory. Our cohort was stratified into two groups, CMD-positive and CMD-negative, with an analysis of the most recent ECG performed prior to the invasive evaluation for each patient.

**RESULTS:** Our cohort comprised 230 patients, of which 60 were classified as CMD-positive and 170 as CMD-negative. T-wave inversion was observed in 30.5% of CMD-positive patients compared to 20% in the CMD-negative group (*P*=0.09), and ST depression was identified in 6.7% of CMD-positive patients compared to 2.4% of CMD-negative patients (*P*=0.12). There were no differences in conduction disorders, such as left bundle branch block (LBBB), left anterior hemiblock (LAHB), left posterior hemiblock (LPHB), right bundle branch block (RBBB), incomplete right bundle branch block (ICRBBB), and bifascicular block, between the two groups. There were no discernible distinctions in the ECG intervals, including the PR, QRS, and QT. There was a trend of increased left ventricular hypertrophy (LVH) according to the ECG criteria in patients with CMD (11.7% vs. 5.3%; *P*=0.09).

**CONCLUSION:** Our study revealed a tendency toward ischemic and hypertrophic ECG changes among CMD-positive patients, but no differences in the occurrence of conduction disorders in patients with and without CMD.

**REGISTRATION:** URL: https://clinicaltrials.gov; Unique Identifier: NCT05960474

**GRAPHIC ABSTRACT:** A graphic abstract is available for this article.

Graphic Abstract.
Clinical pathway for patients with chest pain undergoing ECG and coronary microvascular assessment, highlighting key findings, and insights.

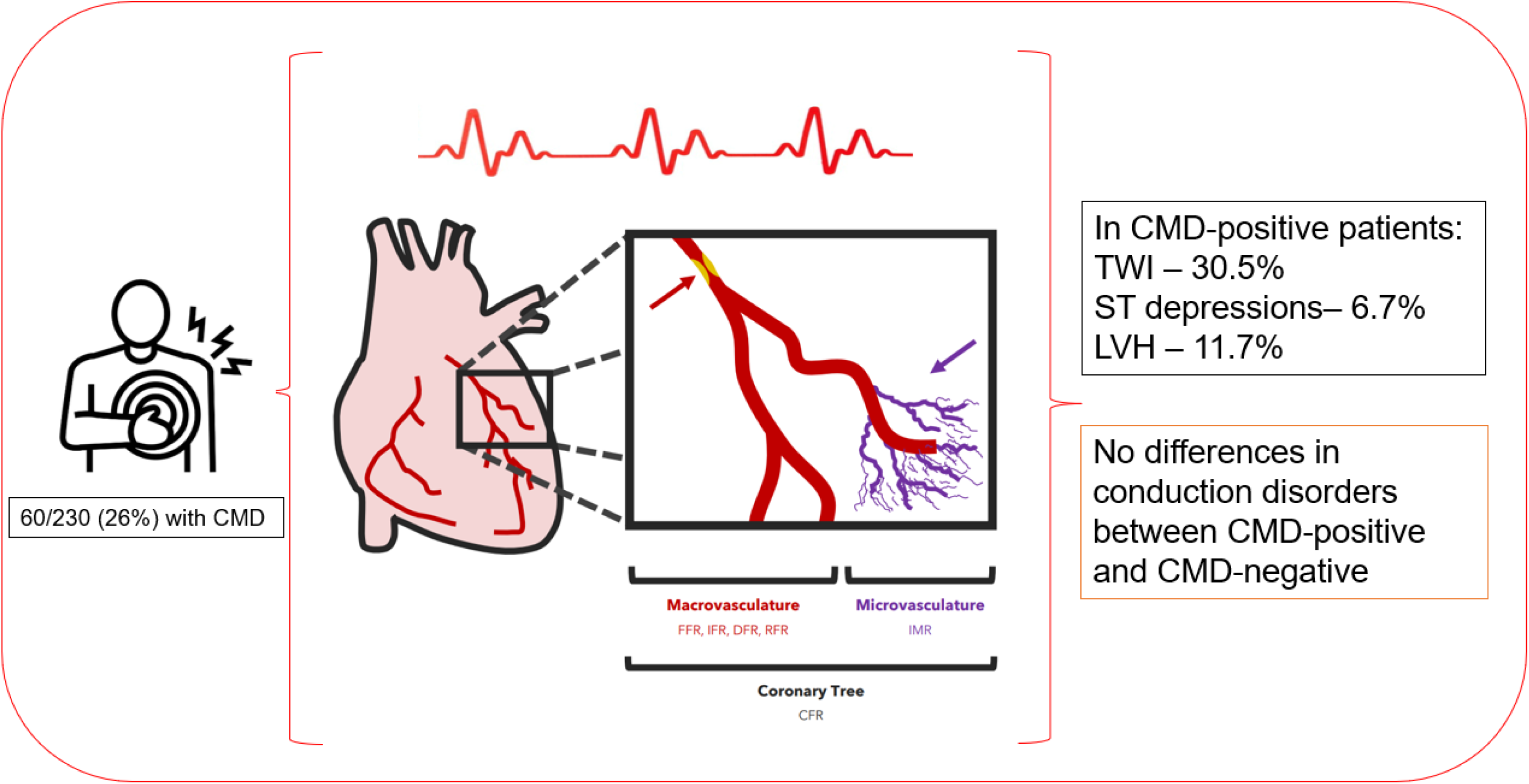

**Clinical Perspectives:** *WHAT IS KNOWN?:*

- Limited research has explored the impact of coronary microvascular dysfunction (CMD) on surface electrocardiogram (ECG) changes.
- The bolus thermodilution method has demonstrated efficacy for the invasive assessment of the coronary microvasculature in patients with symptomatic chest pain. These tools can aid in early detection and diagnosis of CMD.

*WHAT THE STUDY ADDS:*

- In this study from the Coronary Microvascular Disease Registry (CMDR), higher rates of T-wave inversion and ST depression were observed in CMD-positive patients, providing valuable clinical insights. These ECG changes were similar to those observed in patients with stable angina.
- No difference in conduction disorders or ECG intervals was observed, although a greater tendency for left ventricular hypertrophy in CMD-positive patients suggests the importance of monitoring for cardiac structural changes and their potential association with CMD.

## INTRODUCTION

Coronary microvascular dysfunction (CMD), an increasingly recognized cause of cardiac ischemia, poses a significant risk for major adverse cardiovascular events that may impact quality of life^1^. While there is often normal epicardial flow, the effects of disrupted microvascular flow and impaired coronary blood flow have not been fully explored^2^. The electrical conduction system of the heart, such as the sinoatrial node, atrioventricular node, and atrioventricular bundle branch, is also reliant on coronary blood flow^3^. Furthermore, most of the blood supply to the bundle branch is derived from interventricular septal branches originating from the left coronary system. There is a lack of research on whether CMD is associated with conduction disturbances and arrhythmias that can manifest on surface electrocardiograms (ECG).

In cardiovascular evaluation, ECG changes, particularly ST interval and T-wave alterations, are a frequent and fundamental part of noninvasive assessment^4,5^. However, these changes have not received widespread attention or debate in the context of microvascular dysfunction, despite some studies hinting at a link between atrial fibrillation^6^, non-specific ECG alterations, and exercise-induced changes in CMD patients^7^. Furthermore, left ventricular hypertrophy (LVH) can manifest as a physiological response to physical exercise or as a pathological condition, which may be either a primary (genetic) or compensatory response to increased left ventricular (LV) afterload. In individuals with both primary and secondary LVH, there is notable evidence of CMD, which in theory can also be seen in ECG^8^.

Our study aimed to assess CMD and conduction system abnormalities represented by ECG changes. In doing so, we aimed to improve our understanding of the broader impact of on the spectrum of cardiovascular diseases and define the means of noninvasive assessment.

## METHODS

### Establishment of the Coronary Microvascular Disease Registry (CMDR)

The Coronary Microvascular Disease Registry (CMDR) was established to include patients with angina with nonobstructive coronary arteries (ANOCA) who had undergone invasive hemodynamic assessment of the coronary microvasculature^2^. The CMDR is registered as NCT5960474 on clinicaltrials.gov. The data for this registry were obtained from established hospitals, clinical care databases, and existing hospital data collection systems. Notably, no research-specific interventional procedures were performed in this study. Instead, all the procedures were guided by clinical judgment and tailored to the specific needs of each patient. The design and conduct of the registry strictly adhered to the MedStar Health Institutional Review Board (IRB).

### Patient Enrollment and Data Collection

Patients were enrolled at two distinct hospitals: MedStar Washington Hospital Center in Washington, DC, and MedStar Southern Maryland Hospital Center in Clinton, MD, USA. The first patient was enrolled in the registry in August 2021, and data from this patient through November 2023 were included in the current analysis. The CMDR collected a comprehensive array of baseline characteristics, comorbidities, medication details, chest pain severity, results of noninvasive cardiovascular tests, laboratory tests, coronary anatomy from angiography, physiological measurements, and postprocedural outcomes. The registry also included ECG tracings of all patients.

### Assessment of Microvascular Physiology

The CoroVentis CoroFlow Cardiovascular System (Abbott Laboratories, Chicago, IL, USA) was used to assess microvascular physiology. Nonhyperemic resting indices were measured using a physiological pressure wire (PressureWire™ X Guidewire, Abbott Laboratories)^9^. Additionally, indices such as coronary flow reserve (CFR) and the index of microcirculatory resistance (IMR) have been evaluated using thermodilution technology^10,11^. The outcomes of these measurements were methodically recorded and integrated into the CMDR along with the corresponding findings from coronary angiography.

### Patient Classification and Analysis, Inclusion and Exclusion

Patients diagnosed with CMD were classified based on specific criteria: a CFR value of less than 2.0-2.5 and an IMR^12^ equal to or exceeding 25. For precise classification, cases within a borderline range were thoroughly evaluated by a multidisciplinary heart team to ascertain the likelihood of CMD diagnosis^2^. The registry included 246 patients at the time of analysis. However, patients undergoing vasospasm assessment with acetylcholine, those with paced rhythms on their ECG, and those with no ECG tracings in their medical records in the year prior to the invasive assessment were excluded. We also excluded patients who underwent invasive assessment in a vessel other than the LAD, resulting in a final cohort of 230 patients.

### Electrocardiography Assessment

Prior to invasive assessment, the ECGs were examined. Only ECGs conducted within the year before the CMD assessment and interpreted by two cardiologists were considered. Parameters such as PR, QRS, and QT intervals were assessed along with the detection of ischemic changes such as T-wave inversions and ST depressions. Additionally, the criteria for left ventricular hypertrophy (LVH) and axis deviation were evaluated, as well as conduction deficits, fractionated QRS complexes, and premature beats. All analyzed ECGs demonstrated sinus rhythm.

### Statistical Analysis

To compare the characteristics between the two patient groups, we used the Student’s t-test for continuous variables and the χ2 test for categorical variables. Statistical significance was determined at a significance level of *P*< 0.05. The entire data analysis process was conducted using SAS 9.2 (SAS Institute, Cary, NC).

## RESULTS

Our study included a cohort of 230 patients with nonobstructive coronary artery disease who underwent invasive assessment for CMD and met the specified inclusion and exclusion criteria. The overall mean patient age was 61.2±10.9 years and 67.4% were female. Approximately half of the patients (53.5%) were African American. A total of 170 (74%) patients tested negative for CMD, and 60 (26%) tested positive. An in-depth analysis of baseline characteristics, as elaborated in Table 1, revealed that the demographic and comorbidity profiles of CMD-negative and CMD-positive patients were similar. CMD positive patients were slightly older (65.1±10.0 vs. 59.8±10.9 years; *P*=0.001) and had a lower BMI (28.0±5.9 vs. 32.0±6.9 kg/m^2^; *P*<0.001).

**Table 1.**
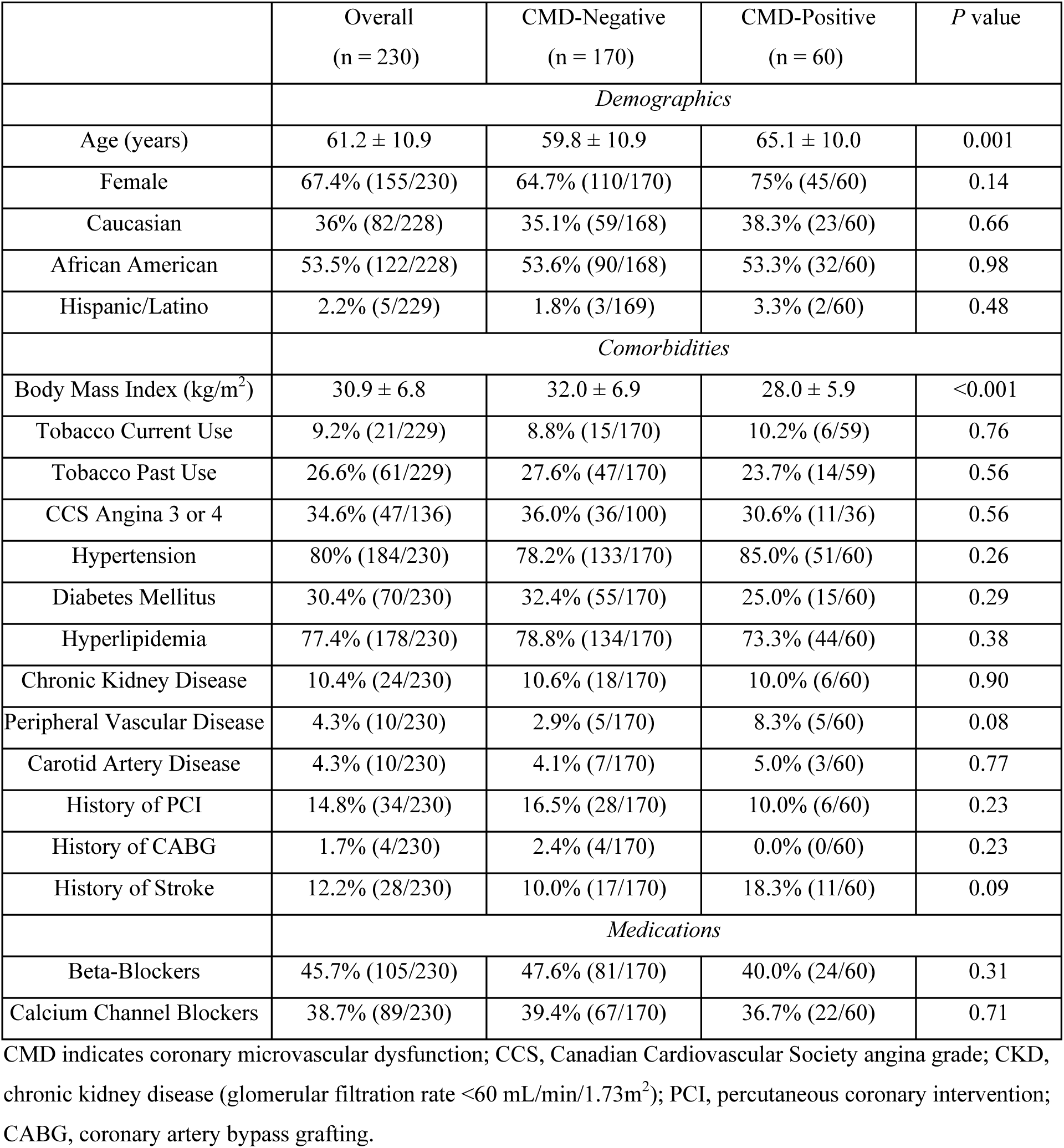
Baseline Characteristics.

Table 2 provides a comprehensive summary of the results obtained from the physiological invasive assessments conducted in the CMD-negative and CMD-positive groups. Table 3 shows the findings derived from ECG assessments. Particularly noteworthy are the observations related to T-wave inversions, which were noted in 30.5% of CMD-positive patients compared to 20% in the CMD-negative group, however, while there was a trend towards CMD positive patients this was not statistically significant (*P*=0.09). Likewise, our study highlights the incidence of ST depression in 6.7% of CMD-positive patients compared to 2.4% among CMD-negative patients (*P*=0.12). Our analysis detected no differences in conduction disorders, including left bundle branch block (LBBB), left anterior hemiblock (LAHB), left posterior hemiblock (LPHB), right bundle branch block (RBBB), incomplete right bundle branch block (ICRBBB), and bifascicular block, between the two groups. Furthermore, no appreciable variances were observed in the ECG intervals, including the PR, QRS, and QT. Our study also identified left ventricular hypertrophy (LVH) using electrocardiographic criteria in 11.7% of CMD-positive patients, whereas this was noted in 5.3% of CMD-negative patients, indicating a trend (*P*=0.09).

**Table 2.** Physiological Assessment.

|  | Overall<br>(n = 230) | CMD-Negative<br>(n = 170) | CMD-Positive<br>(n = 60) | *P* value |
| --- | --- | --- | --- | --- |
| IMR | 18.73 ± 13.35 | 13.08 ± 4.95 | 34.64 ± 16.45 | <.001 |
| CFR | 3.16 ± 1.78 | 3.61 ± 1.82 | 1.92 ± 0.87 | <.001 |
| RFR | 0.88 ± 0.27 | 0.87 ± 0.24 | 0.91 ± 0.35 | 0.42 |
| RRR | 3.61 ± 2.13 | 4.1 ± 2.18 | 2.16 ± 1.05 | <.001 |

CMD indicates coronary microvascular dysfunction; IMR, index of microcirculatory resistance; CFR, coronary flow reserve; RFR, resting full-cycle ratio; RRR, resistive reserve ratio.

**Table 3.** Electrocardiographic Findings.

|  | Overall<br>(n = 230) | CMD-Negative<br>(n = 170) | CMD-Positive<br>(n = 60) | *P* value |
| --- | --- | --- | --- | --- |
| Heart rate (beats/min) | 71.2 ± 12.8 | 71.4 ± 12.9 | 70.6 ± 12.8 | 0.71 |
| PR interval (ms) | 165.7 ± 28.7 | 166.4 ± 28.3 | 163.7 ± 30.0 | 0.54 |
| QRS interval (ms) | 91.4 ± 18.5 | 92.2 ± 18.2 | 89.4 ± 19.2 | 0.31 |
| QT interval (ms) | 408.1 ± 36.7 | 407.6 ± 36.3 | 409.5 ± 38.2 | 0.73 |
| QTc interval (ms) | 438.9 ± 33.7 | 439.1 ± 33.4 | 438.3 ± 34.73 | 0.87 |
| TWI | 22.7% (52/229) | 20.0% (34/170) | 30.5% (18/59) | 0.09 |
| ST depressions | 3.5% (8/229) | 2.4% (4/169) | 6.7% (4/60) | 0.12 |
| Fragmented QRS | 27% (62/230) | 30% (51/170) | 18.3% (11/60) | 0.08 |
| LBBB | 2.6% (6/230) | 2.4% (4/170) | 3.3% (2/60) | 0.68 |
| LAHB | 2.2% (5/230) | 1.8% (3/170) | 3.3% (2/60) | 0.47 |
| LPHB | 0% (0/230) | 0% (0/170) | 0% (0/60) | - |
| RBBB | 2.2% (5/230) | 2.9% (5/170) | 0% (0/60) | 0.18 |
| ICRBBB | 3% (7/230) | 2.9% (5/170) | 3.3% (2/60) | 0.88 |
| Bifasicular block | 0.4% (1/230) | 0.6% (1/170) | 0% (0/60) | 0.55 |
| Left axis deviation | 7% (16/230) | 5.9% (10/170) | 10% (6/60) | 0.28 |
| LVH | 7% (16/229) | 5.3% (9/169) | 11.7% (7/60) | 0.09 |
| RVH | 0% (0/230) | 0% (0/170) | 0% (0/60) | - |
| APBs | 0.9% (2/230) | 1.2% (2/170) | 0% (0/60) | 0.39 |
| VPBs | 6.1% (14/230) | 6.5% (11/170) | 5% (3/60) | 0.68 |

TWI indicates T-wave inversions and do not include non-pathological findings such as a single V1 lead TWI; LBBB, left bundle branch block; LAHB, left anterior hemiblock; LPHB, left posterior hemiblock; RBBB, right bundle branch block; ICRBBB, incomplete right bundle branch block; LVH, left ventricular hypertrophy; RVH, right ventricular hypertrophy; APB, atrial premature beat; VPB, ventricular premature beat.

## DISCUSSION

Our study aimed to investigate the ECG findings in patients with CMD. We observed a trend towards ischemic features and LVH in patients with CMD. Interestingly, we did not find an increased prevalence of conduction or arrhythmia disorders in patients with CMD. These findings shed light on the limitations of noninvasive assessments, such as ECG screening, in identifying individuals at risk of CMD and suggest that ischemia caused by CMD does not harm the conduction system of the heart. However, it is possible that some structural heart diseases, such as LVH, are associated with CMD and can be observed on surface ECG.

Resting ECG is a widely available and cost-effective tool for evaluating myocardial ischemia^13^. However, our study and previously limited published data suggest that resting ECG is often normal in most patients with CMD^14^. Unfortunately, there is no specific diagnostic feature in resting ECG for CMD, making it a test with poor sensitivity for establishing the diagnosis. The absence of ECG changes does not rule out the possibility of CMD, and the presentations of such patients may be diverse. Some patients with CMD exhibit ischemic ECG changes, such as ST depression during treadmill exercise testing or borderline ischemic electrocardiogram findings at rest during chest pain episodes. These observations are consistent with the ECG patterns observed in patients with stable angina^15^.

Sinha et al. recently conducted a prospective study to investigate patients with CMD undergoing exercise electrocardiographic stress testing (EST). A total of 102 patients were included in the study, of whom 65% were women, with a mean age of 60±8 years. Among them, 32 patients developed ischemia during EST (ischemic group), whereas 70 patients did not (non-ischemic group)^16^. Both the groups exhibited similar phenotypes. Ischemia during EST was 100% specific for CMD. This study identified acetylcholine flow reserve as the most robust predictor of ischemia during exercise. When endothelium-independent and endothelium-dependent microvascular dysfunction were used as the reference standard, the false-positive rate of EST decreased to 0%. These results highlight the prominence of ischemic changes on the surface ECG during stress in patients with CMD. Additionally, the findings of this study suggest that the baseline ECG would not differ among these patients.

In terms of conduction disorders, research has shown that ischemia affecting the sinoatrial node, atrioventricular node, and atrioventricular bundle branch is associated with conduction disorders; though, this is more commonly observed in acute events with complete occlusion of epicardial flow^17^. However, patients with CMD exhibit a pattern more reminiscent of that of patients with stable angina who usually experience transient focal or global myocardial ischemia. Typically, ischemic episodes are brief and do not lead to myocardial cell damage. However, prolonged or recurrent chronic ischemia can eventually result in left ventricular dysfunction or arrhythmias that may manifest on surface electrocardiography. This may help explain our findings regarding the lack of association with conduction disorders in patients with CMD.

While our study followed the standard approach to ECG interpretation, we acknowledge the potential for advancements in technology. Artificial intelligence (AI) is now being employed to detect subtle ECG changes, and Ahmad et al. demonstrated the potential of AI in identifying patients at higher risk for conditions such as atrial fibrillation, even in CMD cohorts^18^. Several technologies are currently being evaluated for the use of AI models to non-invasively detect ischemia in ECGs, which may enhance our ability to diagnose CMD in the future^19^.

### Limitations

Our study had some limitations. It was only conducted at two medical centers, was retrospective in nature, the sample size was small, and the diagnosis of CMD relied on non-continuous thermodilution injection. Additionally, not all patients with chest pain were referred for CMD assessment, which could introduce a selection bias. We did not investigate stress ECGs during chest pain episodes and invasive electrophysiological studies were not performed to comprehensively assess conduction.

### Conclusions

In conclusion, our study did not identify significant ECG changes in patients with CMD when compared to patients with similar chest pain without CMD. This suggests that patients with CMD exhibit electrophysiological behavior similar to that of patients with stable angina and do not demonstrate a higher incidence of ECG changes. Our data suggest that abnormal surface ECG findings were not affected by the presence of CMD.

## Data Availability

Data from the manuscript is available upon request.

## Nonstandard Abbreviations and Acronyms

ANOCA: angina with nonobstructive coronary arteries
CFR: coronary flow reserve
CMD: coronary microvascular dysfunction
CMDR: coronary microvascular disease registry
ECG: electrocardiogram
ICRBBB: incomplete right bundle branch block
IMR: index of microcirculatory resistance
LBBB: left bundle branch block
LAHB: left anterior hemiblock
LPHB: left posterior hemiblock
LVH: left ventricular hypertrophy
PPM: permanent pacemaker
RBBB: right bundle branch block
RFR: resting full-cycle ratio
RRR: resistive reserve ratio

## Acknowledgments

None.

## REFERENCES

1. Vancheri F, Longo G, Vancheri S, Henein M. Coronary Microvascular Dysfunction. J Clin Med. 2020;9:2880. doi: 10.3390/jcm9092880

2. Case BC, Merdler I, Medranda GA, Zhang C, Ozturk ST, Sawant V, Margulies AD, Ben-Dor I, Waksman R, Hashim HD. Understanding Patient Characteristics and Coronary Microvasculature: Early Insights from the Coronary Microvascular Disease Registry. Am J Cardiol. 2023;205:97–103. doi: 10.1016/j.amjcard.2023.07.159

3. Futami C, Tanuma K, Tanuma Y, Saito T. The arterial blood supply of the conducting system in normal human hearts. Surg Radiol Anat SRA. 2003;25:42–49. doi: 10.1007/s00276-002-0085-7

4. Pollehn T, Brady WJ, Perron AD, Morris F. The electrocardiographic differential diagnosis of ST segment depression. Emerg Med J. 2002;19:129–135. doi: 10.1136/emj.19.2.129

5. Said SA, Bloo R, de Nooijer R, Slootweg A. Cardiac and non-cardiac causes of T-wave inversion in the precordial leads in adult subjects: A Dutch case series and review of the literature. World J Cardiol. 2015;7:86–100. doi: 10.4330/wjc.v7.i2.86

6. Ozcan C, Allan T, Besser SA, de la Pena A, Blair J. The relationship between coronary microvascular dysfunction, atrial fibrillation and heart failure with preserved ejection fraction. Am J Cardiovasc Dis. 2021;11:29–38.

7. Rahman H, Ryan M, Lumley M, Modi B, McConkey H, Ellis H, Scannell C, Clapp B, Marber M, Webb A, Chiribiri A, et al. Coronary Microvascular Dysfunction Is Associated With Myocardial Ischemia and Abnormal Coronary Perfusion During Exercise. Circulation. 2019;140:1805–1816. doi: 10.1161/CIRCULATIONAHA.119.041595

8. Camici PG, Tschöpe C, Di Carli MF, Rimoldi O, Van Linthout S. Coronary microvascular dysfunction in hypertrophy and heart failure. Cardiovasc Res. 2020;116:806–816. doi: 10.1093/cvr/cvaa023

9. Fearon WF, Kobayashi Y. Invasive Assessment of the Coronary Microvasculature. Circ Cardiovasc Interv. 2017;10:e005361. doi: 10.1161/CIRCINTERVENTIONS.117.005361

10. Keulards DCJ, El Farissi M, Tonino Pim AL, Teeuwen K, Vlaar P-J, van Hagen E, Wijnbergen IF, de Vos A, Brueren GRG, van’t Veer M, Pijls NHJ. Thermodilution-Based Invasive Assessment of Absolute Coronary Blood Flow and Microvascular Resistance: Quantification of Microvascular (Dys)Function? J Intervent Cardiol. 2020;2020:5024971. doi: 10.1155/2020/5024971

11. Medranda GA, Case BC, Hashim H, Rogers T, Bernardo NL, Ben-Dor I, Satler LF, Waksman R. Early Real-World Experience Performing Invasive Hemodynamic Assessment of the Coronary Microvasculature. Cardiovasc Revasc Med. 2022;40:53–54. doi: 10.1016/j.amjcard.2023.07.159

12. Peregrina EF, García-García HM, Roselló JS, Sanchez JS, Kotronias R, Scarsini R, Tebaldi M, Echavarría-Pinto M, De Maria GL. Angiography-Derived Index of Microcirculatory Resistance in the Assessment of Coronary Microvascular Status: A Systematic Review and Meta-Analysis. Cardiovasc Revasc Med. 2022;40:76. doi: 10.1002/ccd.30174

13. Jonas DE, Reddy S, Middleton JC, Barclay C, Green J, Baker C, Asher GN. Screening for Cardiovascular Disease Risk With Resting or Exercise Electrocardiography: Evidence Report and Systematic Review for the US Preventive Services Task Force. JAMA. 2018;319:2315–2328. doi: 10.1001/jama.2018.6897

14. Taqueti VR, Carli MFD. Coronary Microvascular Disease Pathogenic Mechanisms and Therapeutic Options: JACC State-of-the-Art Review. J Am Coll Cardiol. 2018;72:2625. doi: 10.1016/j.jacc.2018.09.042

15. Gulati M, Levy PD, Mukherjee D, Amsterdam E, Bhatt DL, Birtcher KK, Blankstein R, Boyd J, Bullock-Palmer RP, Conejo T, et al. 2021 AHA/ACC/ASE/CHEST/SAEM/SCCT/SCMR Guideline for the Evaluation and Diagnosis of Chest Pain: A Report of the American College of Cardiology/American Heart Association Joint Committee on Clinical Practice Guidelines. Circulation. 2021;144:e368–e454. doi: 10.1161/CIR.0000000000001029

16. Sinha A, Dutta U, Demir OM, De Silva K, Ellis H, Belford S, Ogden M, Li Kam Wa M, Morgan HP, Shah AM, et al. Rethinking False Positive Exercise Electrocardiographic Stress Tests by Assessing Coronary Microvascular Function. J Am Coll Cardiol. 2024;83:291–299. doi:10.1016/j.jacc.2023.10.034

17. Antzelevitch C, Burashnikov A. Overview of Basic Mechanisms of Cardiac Arrhythmia. Card Electrophysiol Clin. 2011;3:23–45. doi: 10.1016/j.ccep.2010.10.012

18. Ahmad A, Corban MT, Toya T, Attia ZI, Noseworthy PA, Lopez-Jimenez F, Cohen MS, Sara JD, Ozcan I, Lerman LO, et al. Coronary Microvascular Dysfunction and the Risk of Atrial Fibrillation From an Artificial Intelligence-Enabled Electrocardiogram. Circ Arrhythm Electrophysiol. 2021;14:e009947. doi: 10.1161/CIRCEP.121.009947

19. Siontis KC, Noseworthy PA, Attia ZI, Friedman PA. Artificial intelligence-enhanced electrocardiography in cardiovascular disease management. Nat Rev Cardiol. 2021;18:465–478. doi: 10.1038/s41569-020-00503-2

